# Physician Agreement and Confidence in Rural Alaska Medevac Decisions: A Cross-Sectional Vignette Study

**DOI:** 10.64898/2026.09.21.26363599

**Authors:** Chelsea Williams, Stephanie Weter, Jeremy Wood, Henry Saul, Brian Rice

## Abstract

**Purpose:** Rural Alaska relies on air ambulances (“medevacs”) to connect remote villages off the road system to higher-level care. Yet medevac decision-making is complicated by limited guidelines, high costs, and delayed transport. We aimed to quantify variability in physician medevac decisions and identify contributing factors.

**Methods:** We conducted a cross-sectional survey of physicians (n=20) within the Alaska Tribal Health System using 20 clinical vignettes representing varying levels of transport complexity. Participants selected transport modality (medevac, commercial, or remain in village) and rated confidence. Interrater agreement was assessed using Gwet’s AC1. A generalized linear mixed model evaluated predictors of medevac use.

**Findings:** Agreement was “almost perfect” for clearly critical cases (“always medevac,” AC1=0.97) but declined substantially for intermediate scenarios (“possible medevac” AC1=0.39; “never medevac” AC1=0.26; “special considerations” AC1=0.12). Confidence strongly correlated with agreement (r=0.83, p<0.001), forming distinct clusters of high-, medium-, and low-certainty decisions. Vignette type was the dominant driver of medevac use (OR 455.5 for “always medevac”), but decisions did not change with experience (OR=0.99, 0.94-1.04). Between-physician variability persisted after adjustment (ICC 3.8%; MOR 1.67), consistent with differences in risk tolerance.

**Conclusions:** Medevac decision-making is highly consistent in certain clinical cases but interrater agreement is limited for intermediate cases. Without a shared gold standard of “correct” medevac decisions, future clinical decision support cannot rely on automated medevac decision making. Efforts to improve care should focus on deconstructing decisions, enhancing outcome feedback, and developing targeted decision support rather than prescriptive decision models.

## Introduction

Rural areas of the United States have higher mortality rates than urban areas, and the gap between them has increased over the last decade.^1^ Emergency care contributes to these unequal outcomes due to significant difficulties in treating critically ill patients and those with emergency care-sensitive conditions (ECSCs).^2^ Contributing to the rural-urban medical outcome discrepancy are the lack of resources to stabilize patients in rural areas, lack of access to specialists and the longer transport times to reach higher levels of care.^3,4,5^ Hospitals and higher-level specialists are also limited and concentrated in large cities.^6^

Alaska faces some of the most significant challenges surrounding rural health, with a landmass larger than California and Texas combined. It is also one of the most rural, with slightly less than half of Alaska’s population residing outside of large cities, described as “remote rural”.^7^ Additionally, over 80% of Alaska’s rural communities are not connected by roads, requiring travel via air or off-road vehicles (i.e., boats and snowmobiles).^8^ ECSCs, as compared to chronic illnesses, represent the greatest contributor to overall excess mortality in these off-road communities.^9^

Most of these rural Alaska communities are small villages composed largely of Alaskan Native peoples living on their ancestral lands. Village medical care is provided by the Alaska Tribal Health System (ATHS) employing a “spoke and hub” system of village clinics (spoke), critical access hospital (regional hub), tertiary referral hospital (central hub). With the rare exception, village clinics are staffed by non-physician clinicians who are either advanced practice providers or Community Health Aides/Practitioners (CHA/Ps). While CHA/P’s serve a critical role, their training is limited and variable, spanning at least two years with a combination of didactics and on the job training.^8^ The EMS system in rural Alaska is largely composed of the village clinic provider, remote physician consultants, and the responding air medevac nurses/paramedics.^10^ For patients in rural Alaska, shared decision making – often between a CHA/P on scene and a remote physician - is used to determine the necessity of activating a medevac. While essential to the chain of survival in rural Alaska, the cost of an air medevac contributes significantly to the cost of healthcare within the state. A 2017 national report listed a median charge of $10,199 for air medevacs nationwide.^11^ The cost of an air medevac in Alaska can exceed $100,000 due to longer travel distances and lack of available specialists sometimes necessitating transport out of state.^12^ Furthermore, medevac resources are very limited and responses are delayed.

The median response time in rural Alaska between activating a medevac and getting the patient to a receiving hospital is approximately four hours, and response times can sometimes be greater than 24 hours.^13^ Finally, each region has a finite number of medevac planes and crews available to them, often just one. Frequently using a medevac for one emergency means that it will be unavailable for other emergencies in the region.

Considering the cost and drain on available resources with each air medevac activation, it is essential that providers identify patient conditions that warrant their use. There are few formal guidelines and minimal literature has meaningfully addressed the decision-making process behind these choices. Consensus across the American College of Emergency Physicians (ACEP) and aeromedical services have been published suggesting specific aeromedical care for less common and clearly high risk conditions such as ST-elevation MI, stroke, cardiac arrest and significant trauma.^14^ An internally and externally validated national trauma triage protocol has been developed, helping to guide transfer in patients meeting specific criteria.^15,16^

However, without clear established indications for transfer outside of trauma, this puts complicated and high risk decisions firmly on the shoulders of rural providers who have limited evidence-based medicine to support their choices. Hogarth described these types of decisions as occurring in “wicked” learning environments, defined as those wherein success and failure are ambiguous, feedback is limited, signals are noisy, and the relationship between decisions and outcomes is difficult to infer.^17^ As compared to “kind” environments, where consistent feedback supports learning, physicians in a “wicked” environment are making high-stakes transport decisions with limited ability to observe downstream outcomes, which impairs their ability to improve or adapt their decision making.

This study is a first step in assessing on-the-ground physician consensus in air medevac necessity. Our objective for this study is to identify how much variability exists amongst physicians in medevac decision making, and what contributes to that variability. To obtain that objective, we studied decision making by physicians within the ATHS who are regularly tasked with activating air medevacs by giving them clinical vignettes designed to mirror real weather, logistic and patient characteristics that may impact decision making.

## Methods

### Setting

We conducted a cross-sectional survey of physicians within the Alaska Native Medical Health Consortium (ANTHC) between March and October 2025. All physicians working consistently in the emergency departments at the central hub hospital Alaska Native Medical Center (ANMC), and a regional hub hospital, Maniilaq Health Center (MHC) were eligible to participate. MHC is located in the Northwest Arctic Bureau, a nonmetropolitan region delineated by the U.S. Office of Management and Budget.^18^ Eligible participants were sent an email introducing the study and requesting voluntary participation. The survey was developed and administered in a web-based format with study data collected and managed using REDCap electronic data capture tools hosted at ANTHC.

### Survey and Vignette Development

The survey was separated into two sections, starting with questions about demographics including years in practice, degree achieved, and those surrounding rural experience (location of practice, years employed in rural setting). The second portion of the survey included 20 clinical vignettes with prompts asking participants to choose between different transport options for further care-medical evacuation immediately (“medevac”), commercial airline transfer next available (“commercial”), and no transfer with continued care in village if necessary (“remain in village”). Participants were also asked to grade their level of confidence in their decision for each vignette on a scale of 1 to 10. Vignettes were developed by two physicians with clinical expertise in rural Alaskan emergency care (CW, JW). Vignettes were intended to range in difficulty from easy to challenging, and ranged across four types: 1) “never medevac” – vignettes where medevac was the least appropriate choice (as compared to commercial or remain in village), 2) “always medevac” – vignettes where medevac is the most appropriate choice, 3) “Possible medevac” – medevac is a reasonable choice, but so are commercial and/or remain in village, 4) “special considerations” – medevac or remain in village only reasonable options (not commercial) for unique situations (e.g., traumatic arrest) where emergency transport is not reasonable due to the multiple hours required for a medevac to effect transport (see Supplemental Index 1).

### Data Analysis

Our data represented repeated measures as each physician responded to 20 vignettes and our analysis reflected the need to understand variation both amongst physicians and between vignettes. We reported descriptive statistics about physician demographics and training, and reported agreement amongst physicians across each vignette, and all four vignette types. We defined agreement as whether a physician’s choice matched any prespecified “correct” option for that vignette and computed the Pearson correlation between agreement and decision-level confidence (treated as continuous), calculating the correlation coefficient and two-sided p-value. We used Gwet’s AC1 to quantify interrater agreement because it provides a chance-corrected measure that is more stable than kappa when category frequencies are intentionally unbalanced. We then used a mixed-effects logistic regression model to account for the nested structure of our data: multiple responses from each physician across multiple vignettes. The generalized linear mixed model (GLMM) used “decision to medevac” as our outcome variable, included both physicians and vignettes as random intercepts, and included vignette type, case-specific confidence, experience, and practice location (rural vs. referral hub) as fixed effects. We assessed model performance using variance partitioning, R², and standard prediction metrics (ROC AUC, Brier score, calibration).

Odds ratios (OR) were used to report on fixed effects, and median odds ratio (MOR) and intraclass correlation (ICC) were used to report the random effects in the model. The threshold for significance was set at 0.05, Python was used for all data management, and R was used for data analysis and to produce figures.

### Ethics

The study was approved by Alaska Area Institutional Review Board (IRB), with an IRB reliance agreement with Stanford University, as well as Tribal approval from Alaska Native Tribal Health Consortium, Southcentral Foundation Board of Directors, and Maniilaq Association. Informed consent was obtained from all participants.

## Results

Of the 36 physicians approached for the study, 20 consented to participate (55% response rate), and all 20 completed demographic information and provided responses to all 20 vignettes (n=400/400, 100% completion rate). Respondents’ years in practice median was 8 [IQR 4-13, range 1-30] with 60% having rural experience. The classification of vignettes as well as descriptive statistics of responses can be found in *Table 1*.

**Table 1:** Sample Characteristics and Study Design.

| Characteristic | Value |
| --- | --- |
| **\*\*Physicians\*\*** |  |
| Total Physicians | 20 |
| **\*\*Years in Practice\*\*** |  |
| Median [IQR] | 8 [4 - 13] |
| Range | 1 - 30 years |
| **\*\*Practice Location\*\*** |  |
| Critical Access Hospital | 7 (35%) |
| Tertiary Referral Hospital | 8 (40%) |
| Both | 5 (25%) |
| **\*\*Vignettes\*\*** |  |
| Total Vignettes | 20 |
| Never Medevac | 4 |
| Possible Medevac | 9 |
| Special Considerations | 3 |
| Always Medevac | 4 |
| **\*\*Responses\*\*** |  |
| Total Responses | 400 |
| Appropriate Response to Always Medevac | 98.8% (n=79) |
| Appropriate Response to Never Medevac | 88.8% (n=71) |
| **\*\*Inter-Rater Agreement by Vignette\*\*** |  |
| Median [IQR] | 75.0% [65.0% - 83.8%] |
| Range | 45.0% - 100.0% |
| **\*\*Confidence (1-10 scale)\*\*** |  |
| Median [IQR] | 8 [6 - 10] |
| Range | 1 - 10 |

Participants were largely confident in their decision making with a median score of 8/10 [IQR 6-10] although there was a full range of responses from 1-10.

*Table 2* shows the detailed breakdown of provider decisions categorized by the anticipated response (“always medevac”, “possible medevac”, “never medevac”, “special considerations”) for each vignette, as well as the crude agreement, median confidence for each vignette. We see that the inter-rater agreement is “almost perfect” (AC1= 0.97 [0.92-1.00]) for the “always medevac” category but is only “fair” for both “possible medevac” (AC1 = 0.39 [0.23-0.54]) and “never medevac” (AC1 = 0.26 [-0.03-0.55]), and only “slight” for “special considerations” (AC1 = 0.12 [-0.05-0.29]).

**Table 2:**
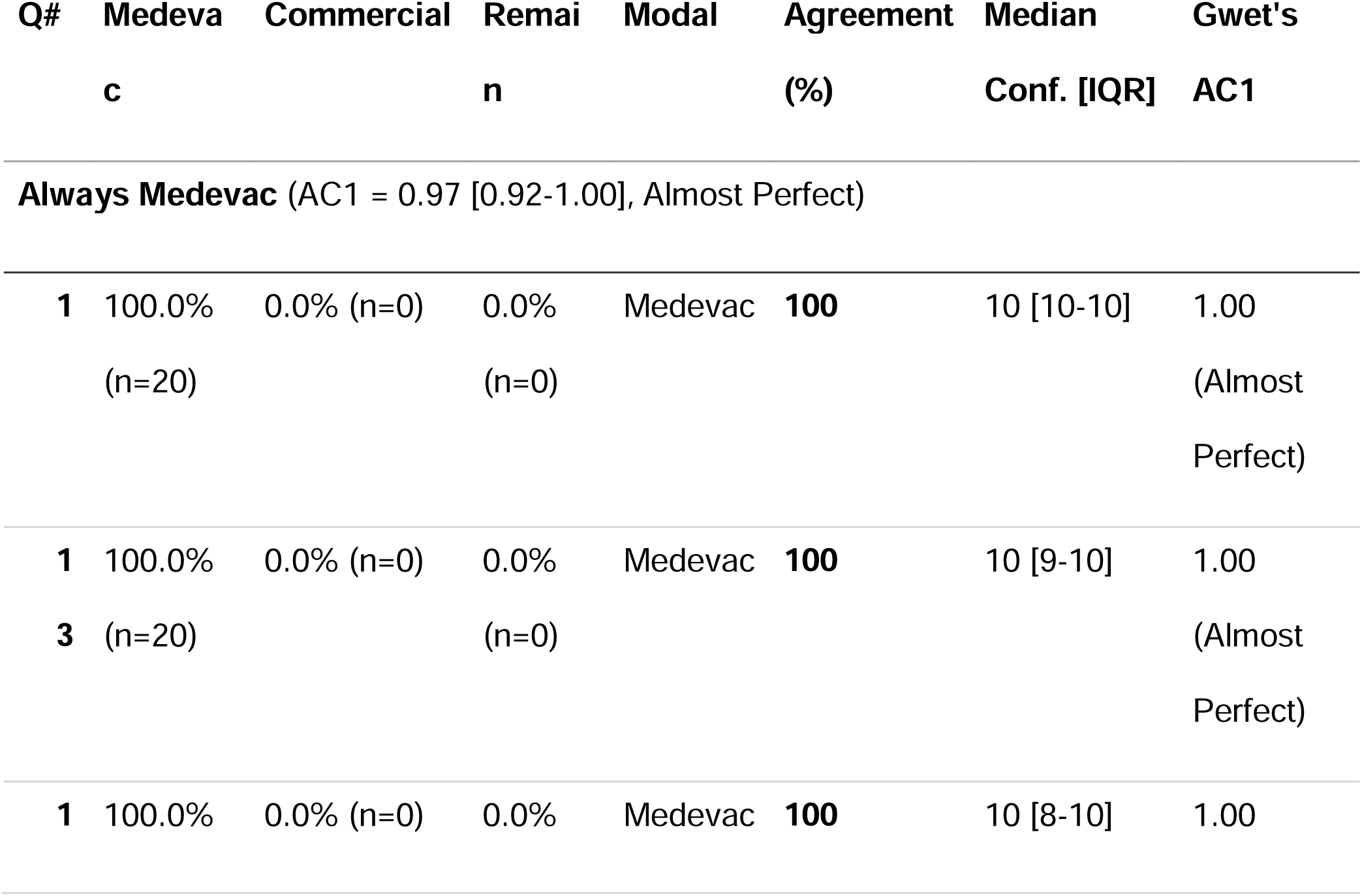
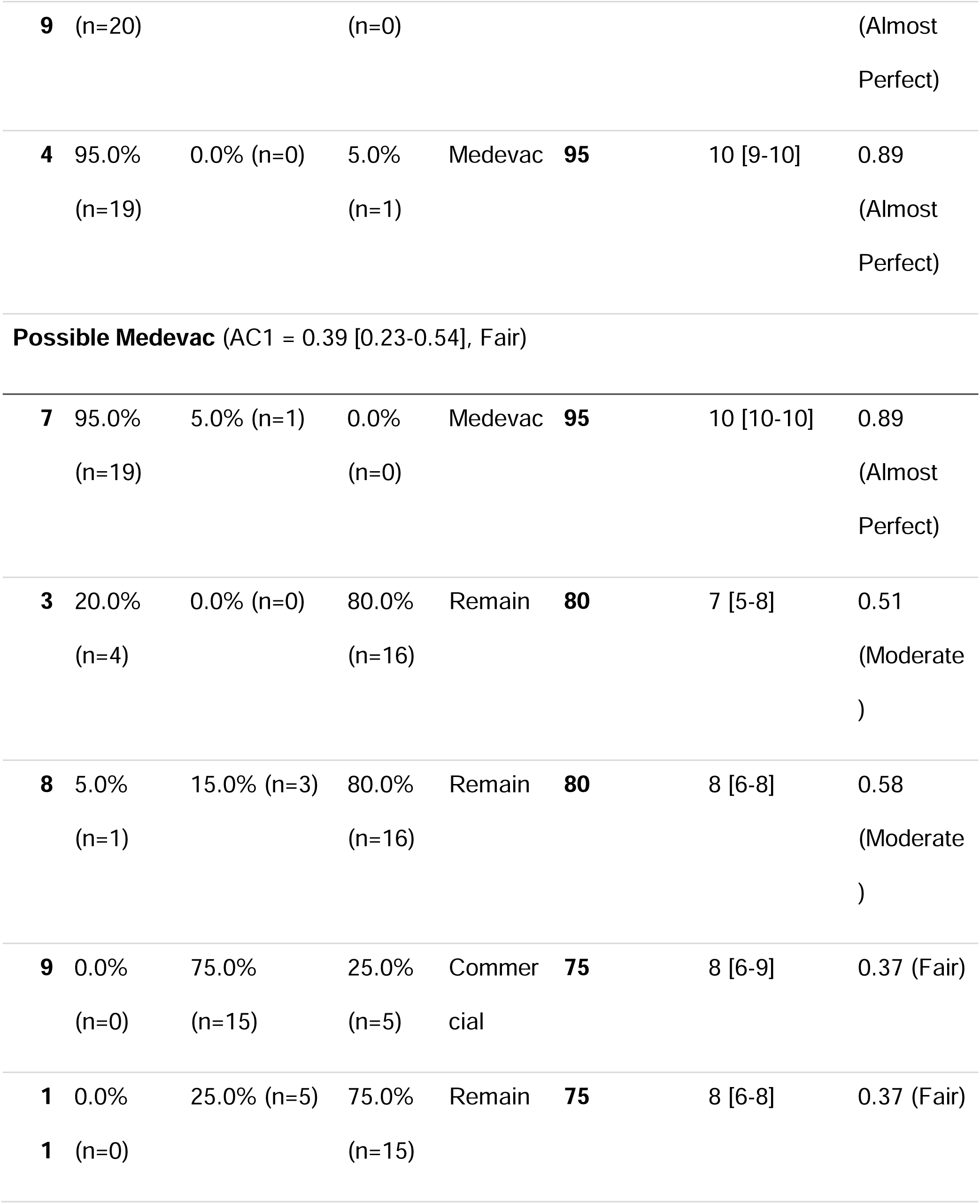
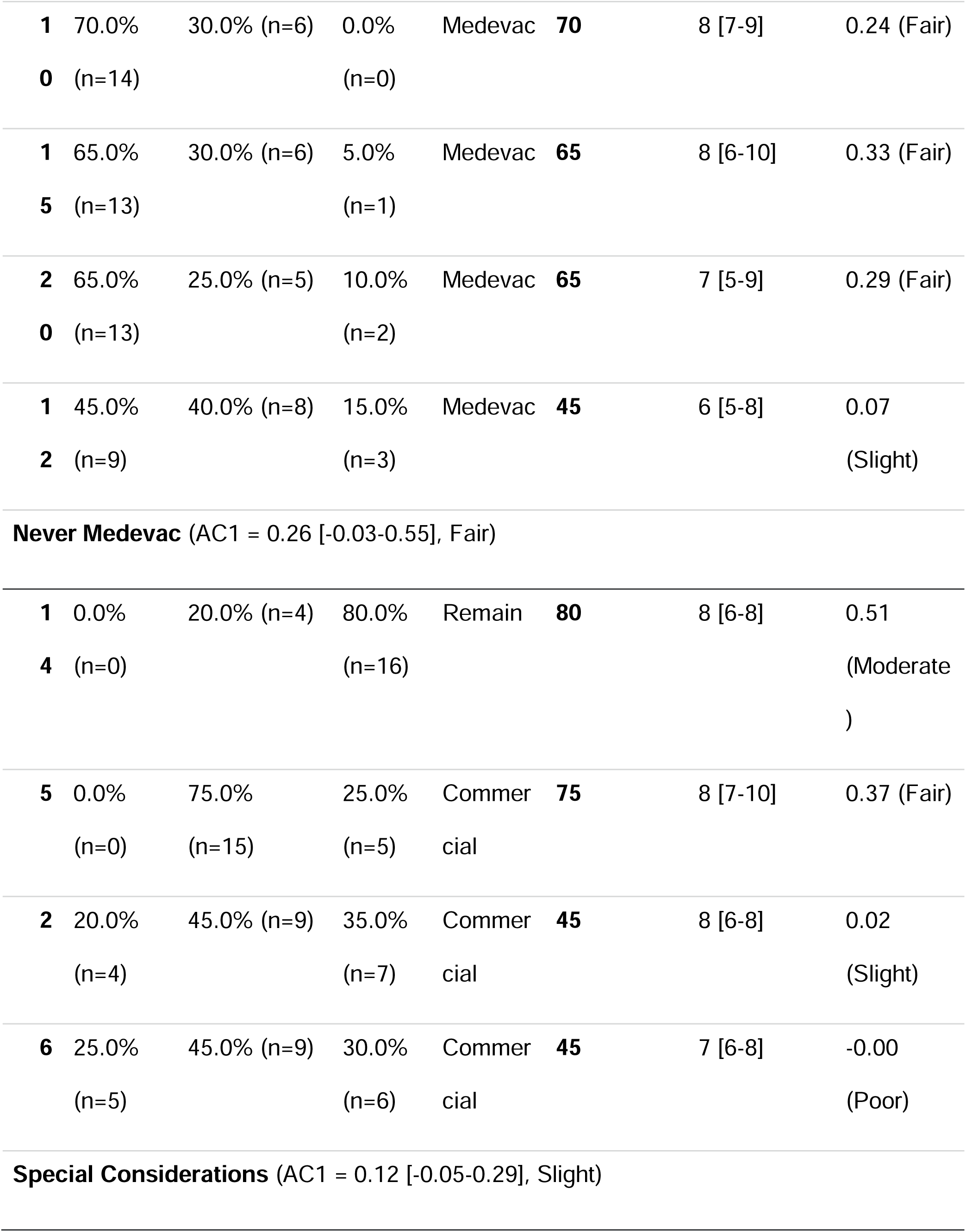
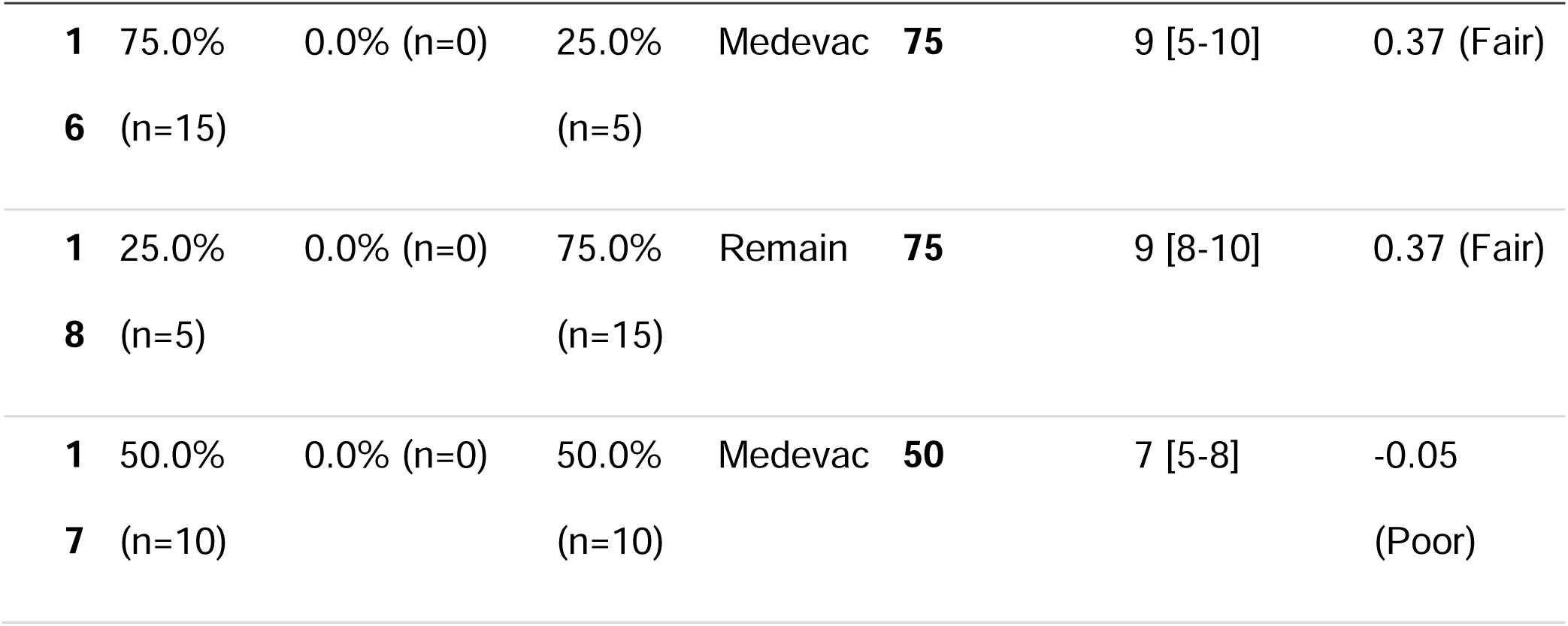
Interrater agreement about medevac vignettes.

We visualized the significant positive correlation (r=0.83, p = <0.001) between agreement and confidence in medevac decision making in Figure 1. This association between increasing confidence and increasing agreement produced three discrete clusters of decisions correlating roughly with “low”, “medium”, and “high” agreement.

**Figure 1:**
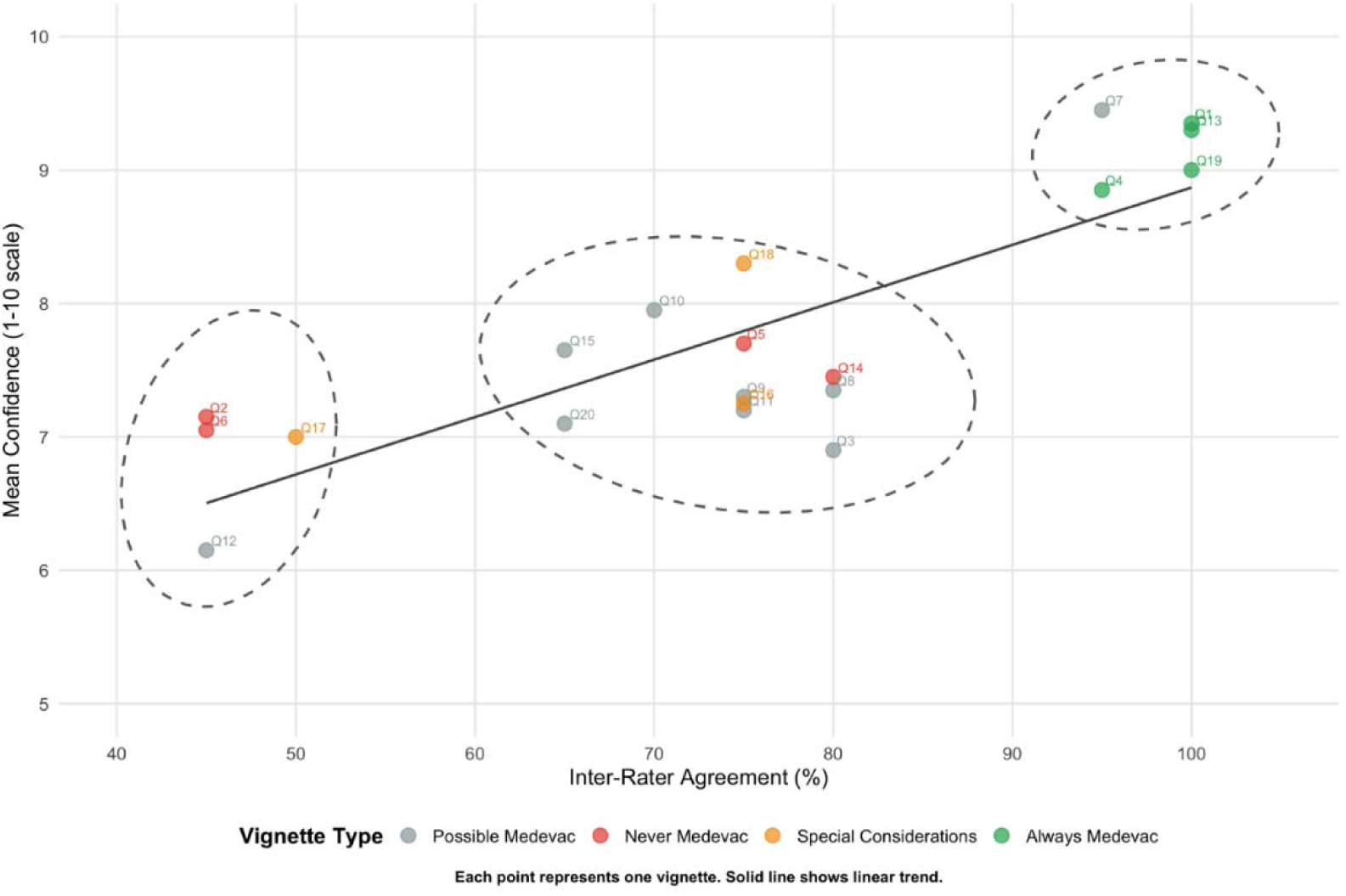
Confidence and Agreement by Vignette

The results of our GLMM (with choosing “medevac” as the outcome variable) are displayed in two tables, one with the fixed effects (Table 3) and one with the variable effects (Table 4). Vignette type had a very large impact with “always medevac” having a large positive association (OR=455.53, [13.40 - 15481.30], p<0.001) as compared to “possible medevac” and “special considerations” vignettes, while “never medevac” vignettes showed a strong inverse association (OR=0.07, [0.00 - 1.05], p<0.055) that narrowly missed conventional statistical significance. Rural experience and level of confidence (as compared to each physician’s baseline) were significantly associated with choosing medevac, while experience did not increase or decrease choosing medevacs.

**Table 3:**
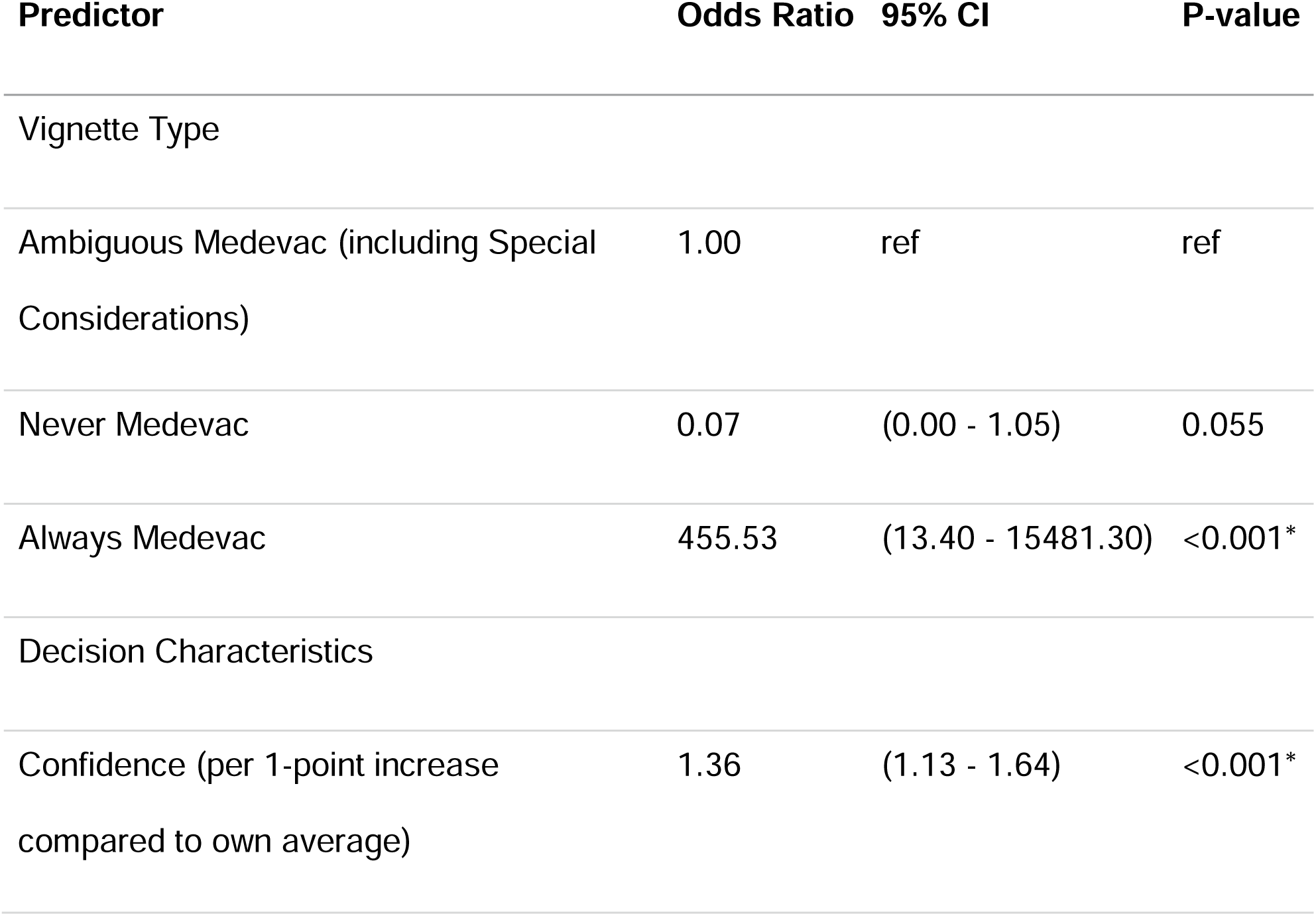
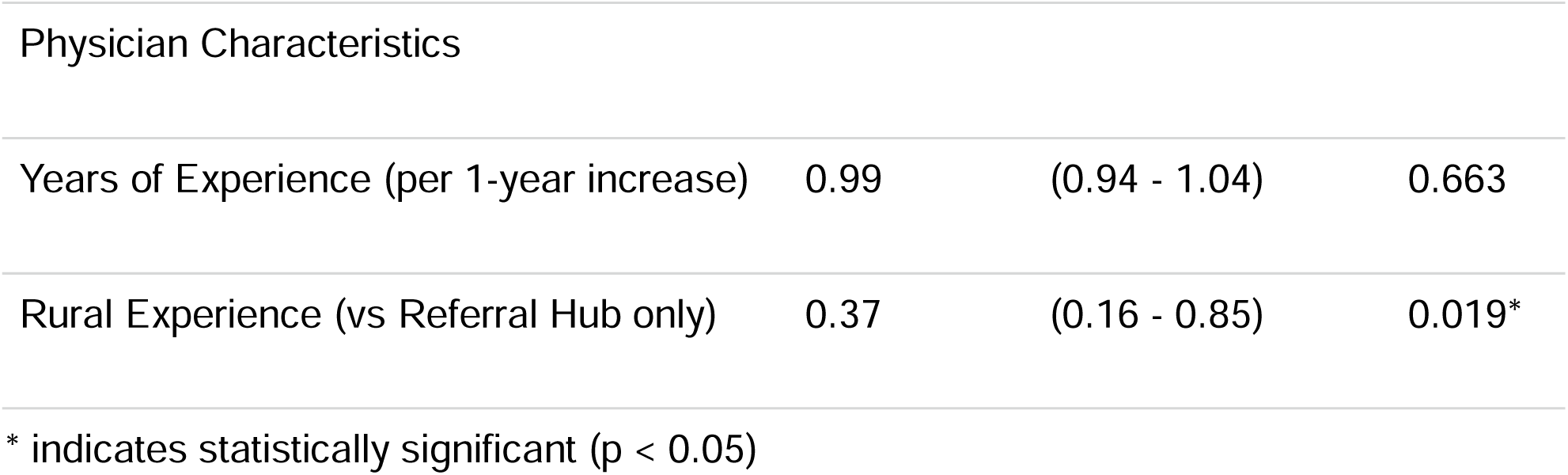
Model Results - All Fixed Effects.

| Predictor | Odds Ratio | 95% CI | P-value |
| --- | --- | --- | --- |
| Vignette Type |  |  |  |
| Ambiguous Medevac (including Special Considerations) | 1.00 | ref | ref |
| Never Medevac | 0.07 | (0.00 - 1.05) | 0.055 |
| Always Medevac | 455.53 | (13.40 - 15481.30) | <0.001* |
| Decision Characteristics |  |  |  |
| Confidence (per 1-point increase compared to own average) | 1.36 | (1.13 - 1.64) | <0.001* |
| Physician Characteristics |  |  |  |
| Years of Experience (per 1-year increase) | 0.99 | (0.94 - 1.04) | 0.663 |
| Rural Experience (vs Referral Hub only) | 0.37 | (0.16 - 0.85) | 0.019* |

\* indicates statistically significant ( $p < 0.05$ )

**Table 4:** Model Results - Variance Components.

| Component | Estimate |
| --- | --- |
| Random Effect Variances |  |
| Physician (log-odds scale) | 0.288 |
| Vignette (log-odds scale) | 4.101 |
| Intraclass Correlations |  |
| ICC (Physician) | 3.8% |
| ICC (Vignette) | 53.4% |
| Median Odds Ratios |  |
| MOR (Physician) | 1.67 |
| MOR (Vignette) | 6.90 |

After adjustment for vignette type, confidence, years of experience, and rural experience, the residual variability was substantial between vignettes (ICC 53.4%; MOR 6.90) but modest between physicians (ICC 3.8%; MOR 1.67) (Table 4).

Marginal R^2^ was 0.57, indicating that the fixed effects accounted for a substantial proportion of variation in medevac decisions. Conditional R^2^ was 0.82, indicating that the full model, including physician- and vignette-level random effects, accounted for substantially more variation.

## Discussion

The above analysis is the first known published investigation of rural medevac provider decision-making. Our vignettes were designed to mimic decision-making in a limited information state with complex logistical and clinical interactions. This experimental design led to several interesting findings. First, there was almost perfect interrater agreement for “always medevac” vignettes (AC1= 0.97 [0.92-1.00]). This suggests that there is a shared understanding of certain patient characteristics that more clearly categorizes them as critical or requiring a higher level of care. In vignette 1 for example, which has 100% agreement among providers, this is likely due to low Glasgow coma score in the setting of high mechanism trauma. This fits with general consensus and evidence-based trauma triage scores.^14,15,16^

The second finding in the other groups of medevac vignettes, where choosing medevac was not the “correct” choice, there was only fair or slight interrater agreement. This supports the anecdotal experience of providers in these regions, who see highly variable decisions made by different physicians based on individual experiences and risk tolerances in all but clear-cut cases. This has profound implications for the potential of developing clinical decision support systems for medevac decision making. Since experienced physicians are unable to agree on the “correct” mode of transport for a borderline patient, it is not feasible for even an advanced model to identify “correct” decisions as that is not a shared “gold standard” label between human experts. This suggests that future work in this space may need to look at supporting individual parts of the medevac decision (e.g. how likely is this patient to escalate to needing ICU-level care if they have to wait in village for more than 12 hours) rather than a holistic “does this patient need a medevac” approach.

Looking at the correlation between agreement and confidence (Figure 1) we see again that providers are more confident and agree more when choosing to medevac for “always medevac” vignettes. This fits with existing literature showing that resource utilization is often driven by both uncertainty and patient acuity.^19^ However, there is an interesting effect seen where three clear clusters of “high”, “medium” and “low” agreement and certainty emerged. This was not an intended part of our experimental design. This novel finding suggests a direction for future research. If we are able to identify which clinical, systems, and environmental features separate these clusters, we may be able to adapt clinical decision support which explicitly addresses those factors.

Several vignettes were designed to incorporate decision making including unique rural logistical challenges. There are certain scenarios which would represent a maximal lights and sirens EMS response on the road system, but which would receive a black tag triage in a remote village due to requiring several hours for a medevac. Vignette 18 presented one such case, a pulseless gunshot wound to the chest. This vignette was designed to represent almost certain mortality, as pulseless traumatic arrest has low survivability in even the highest level of care environments with transport times less than 10 minutes. Transport time of several hours for medevacs would be universally unsurvivable. Yet 5 of the 15 respondents chose to activate a medevac. This suggests that clear rulesets or protocols for medevac guidance may help physicians working within the system who struggle to comprehend the harsh realities of life off of the road system.

Looking at the fixed effects in our GLMM model we identified three main trends-vignette types were the predominant factor driving medevac decisions, rural experience reduced medevac utilization, and years of clinical experience did not affect medevac choices.

The first finding was expected and matched our a priori assumption which informed the experimental design of this vignette-based design. Secondly, we noticed that experience working in the rural critical access hospital reduced medevac utilization. This may be due to non-medevac choices on the “black tags” special cases, or a different overall comfort with less rapid transport. This suggests that incorporating the clinical experience of rural physicians is critical for improving health decisions regarding medevac. Finally, we saw that years of physician experience neither increased nor decreased utilization rates. Previous research has shown that experience in emergency medicine increases risk-tolerance making providers more accepting of uncertainty, utilizing less imaging resources.^20,21^ It is not possible from our study design to evaluate why medevac decisions do not change similarly with age. However, it may be because rural medevac utilization decisions are made in these “wicked” environments.^22^ There is extreme uncertainty in a remote assessment of a patient along with a total absence of feedback. The authorizing provider is almost never the provider who assesses the patient in person and receives no follow-up information about clinical outcomes. The implications here apply to many complex decision-making domains for physicians and suggest that finding ways to provide patient and systems-level feedback to decision makers can help improve quality and efficiency. These issues are especially relevant in rural health systems which are often more resource constrained than their urban counterparts.

Even when controlling for the vignette type, confidence, years of experience, rural experience, and substantial between-vignettes variability, a small but significant between-provider variability was identified (ICC = 3.8%, MOR = 1.67). This means that the odds of some providers ordering a medevac are 67% higher than a theoretical “median” physician. One interpretation of this number is it represents the variable “risk tolerance” that exists between our study physicians in medevac decisions. Being able to isolate and quantify this variable suggests that the widespread clinician impression that disparate risk tolerance drives medevac decision making is likely based in fact. It also suggests that reducing this variability is one route for improving medevac systems quality.

Our study has several limitations. We had to develop our own rural medevac vignettes as no externally validated instruments exist. Additionally, the physicians and systems studied are all based in Alaska. While the implications about rural emergency medicine decision-making are likely externally valid in other places, the lack of a road system and reliance on medevacs in Alaska is a system that is structurally unlike other rural regions.

## Conclusions

Medevac decision-making in rural Alaska is highly reliable in extreme cases but fundamentally indeterminate for a large intermediate group of patients, where neither clinical acuity nor logistics alone define the optimal transport choice. This variability is not simply noise, but a structural feature of a low-feedback, high-uncertainty decision environment lacking a shared gold standard. The absence of consensus among experienced physicians challenges the feasibility of prescriptive guidelines or end-to-end decision automation. Instead, progress will require reframing medevac decisions as composable processes, improving outcome-linked feedback, and developing targeted decision support that addresses specific components of uncertainty. This paradigm has implications for complex decision-making across resource-constrained healthcare systems.

## Data Availability

Data is not available due to the presence of identifiable information

## Acknowledgements

None

## Notes

**Funding sources:** The author(s) declare that financial support was received for the research and/or publication of this article. The research described above were supported by grant K08MD016445 from the National Institute of Minority Health and Health Disparities, grant 3OT2OD032581-01S5-800 from the AIM-AHEAD Artificial Intelligence/Machine Learning Consortium to Advance Health Equity and Researcher Diversity Program. The content is solely the responsibility of the authors and does not necessarily represent the official views of the National Institutes of Health.

### Competing Interest Statement

The authors have declared no competing interest.

